# Potentially inappropriate prescribing of DOACs to people with mechanical heart valves: a federated analysis of 57.9 million patients’ primary care records in situ using OpenSAFELY

**DOI:** 10.1101/2021.07.27.21261136

**Authors:** The OpenSAFELY Collaborative, Louis Fisher, Victoria Speed, Helen J Curtis, Christopher T Rentsch, Angel YS Wong, Anna Schultze, Jon Massey, Peter Inglesby, Caroline E Morton, Marion Wood, Alex J Walker, Jessica Morley, Amir Mehrkar, Seb Bacon, George Hickman, Chris Bates, Richard Croker, David Evans, Tom Ward, Jonathan Cockburn, Simon Davy, Krishnan Bhaskaran, Becky Smith, Elizabeth Williamson, William Hulme, Amelia Green, Rosalind M Eggo, Harriet Forbes, John Tazare, John Parry, Frank Hester, Sam Harper, Jonathan Meadows, Shaun O’Hanlon, Alex Eavis, Richard Jarvis, Dima Avramov, Paul Griffiths, Aaron Fowles, Nasreen Parkes, Ian J Douglas, Stephen JW Evans, Laurie Tomlinson, Brian MacKenna, Liam Smeeth, Ben Goldacre

## Abstract

National guidance was issued during the COVID-19 pandemic to switch patients on warfarin to direct oral anticoagulants (DOACs) where appropriate as these require less frequent blood testing. DOACs are not recommended for patients with mechanical heart valves. We conducted a retrospective cohort study of DOAC prescribing in people with a record of a mechanical heart valve between September 2019 and May 2021, and describe the characteristics of this population. We identified 15,457 individuals with a mechanical heart valve recorded in their records, of whom 1058 (6.8%) had been prescribed a DOAC during the study period. 767 individuals with a record of a mechanical heart valve were currently prescribed a DOAC as of May 31st 2021. This is suggestive of inappropriate prescribing of DOACs in individuals with mechanical heart valves. Direct alerts have been issued to clinicians through their EHR software informing the issue. We show that the OpenSAFELY platform can be used for rapid audit and feedback to mitigate the indirect health impacts of COVID-19 on the NHS. We will monitor changes in prescribing for this risk group over the following months.

## Introduction

NHS England issued guidance for anticoagulant services during the COVID-19 pandemic including advice on the safe switching of patients on warfarin to direct oral anticoagulants (DOACs), where appropriate^1^. As DOACs require less frequent blood testing, this supported the reduction of pressures being experienced by the NHS and the lowering of the risk of virus transmission. We have previously shown that this guidance was followed by increased switching of anticoagulants to DOACs^2^.

The use of DOACs in people with mechanical heart valves is not recommended and was explicitly advised against in the guidance issued during the switching programme^1,3^. Despite this, the national patient safety team at NHS England received anecdotal reports of individuals with mechanical heart valves being inappropriately prescribed a DOAC as a result of the switching programme. A national audit would previously have been challenging on an issue such as this: however the establishment of the OpenSAFELY analytics platform for research and service improvement during COVID-19 raised the possibility of conducting a detailed analysis of adherence and breaches using the full raw pseudonymised linked electronic health records of almost the whole population.

We therefore set out to describe prescriptions of DOACs to people with a record of mechanical heart valves throughout the COVID-19 pandemic in England, using OpenSAFELY.

## Methods

Working on behalf of NHS England, we used the OpenSAFELY framework to conduct a retrospective cohort study of DOAC prescribing to people with a record of mechanical heart valves across the full pseudonymised patient primary care records for 57.9 million people (97%) registered at a general practice in England using either TPP or EMIS software

Between September 2019 and May 2021, at the beginning of each month, we identified individuals aged 16 or older who had a SNOMED-CT code selected as being explicitly indicative of ever receiving a mechanical heart valve^4^. We then ascertained the number of people who were also prescribed a DOAC in each month^5^. People were classified as currently prescribed a DOAC if they had a DOAC prescription between March 1st 2021 and May 31st 2021. We describe the following characteristics of this population; age band, sex, ethnicity, quintile of Index of Multiple Deprivation (IMD), record of previous atrial fibrillation (AF) and specific mechanical valve code. All analytical code, results and clinical codelists used in this study are openly available for inspection and re-use at reports.opensafely.org.

## Results

We identified 15,457 people in the study cohort coded as having a mechanical valve at any time before the start of May 2021. Of these, 1,058 (6.8%) were identified as having been prescribed a DOAC between September 2019 and May 2021 of which 768 were currently prescribed a DOAC as of May 31st 2021.

Among people coded to have a mechanical valve and currently prescribed a DOAC (Table 1), 62% were male and 38% were female. 67% were of white ethnicity, 4% were non-white and 29% did not have a recorded ethnicity. The most common codes were “*mechanical prosthetic aortic valve replacement - 174929002*” (n= 538) and “*mechanical prosthetic mitral valve replacement - 431339008*” (n = 187). 85% had a previously recorded AF diagnosis. In practices using EMIS software, 7.6% of people coded as having a mechanical valve were prescribed a DOAC, vs 3.2% in practices using TPP software.

**Table 1.** Counts of people with mechanical valves currently prescribed a DOAC by demographic characteristics between March 2021 and May 2021.

| Characteristic |  | Count of individuals coded as having a mechanical valve prescribed a DOAC (column % within category) | Population coded as having a mechanical valve (column % within category) |
| --- | --- | --- | --- |
| Total |  | 768 (100.0) | 13,777 (100.0) |
| Sex | Male | 473 (61.6) | 8,847 (64.2) |
|  | Female | 295 (38.4) | 4,930 (35.8) |
| Ethnicity | White | 516 (67.2) | 8,427 (61.2) |
|  | Non-white | 32 (4.2) | 1,177 (8.5) |
|  | Unknown | 220 (28.6) | 4,173 (30.3) |
| Age Band | 16-49 | 22 (2.9) | 2,087 (15.1) |
|  | 50-59 | 29 (3.8) | 2,362 (17.1) |
|  | 60-69 | 74 (9.6) | 3,041 (22.1) |
|  | 70-79 | 271 (35.3) | 3,720 (27.0) |
|  | 80+ | 372 (48.4) | 2,567 (18.6) |
| IMD | Most deprived or Missing | 97 (12.6) | 2,560 (18.6) |
|  | 2 | 126 (16.4) | 2,677 (19.4) |
|  | 3 | 162 (21.1) | 2,956 (21.5) |
|  | 4 | 177 (23.0) | 2,846 (20.7) |
|  | Least deprived | 206 (26.8) | 2,738 (19.9) |
| Mechanical valve code | Mechanical prosthetic aortic valve replacement (174929002) | 538 (70.1) | 9,660 (70.1) |
|  | Mechanical prosthetic mitral valve replacement (431339008) | 187 (24.3) | 3,297 (23.9) |
|  | Other | 43 (5.6) | 820 (6.0) |
| Record of previous atrial fibrillation | Yes | 655 (85.3) | 4,913 (35.7) |
|  | No | 113 (14.7) | 8,864 (64.3) |
| EHR Software Vendor | TPP | 197 (25.7) | 6,247 (45.3) |
|  | EMIS | 571 (74.3) | 7,530 (54.7) |

We observed a progressive increase in the monthly number of people with mechanical valves prescribed DOACs, with an increase from 472 (35.8 per 1000 patients coded as having a mechanical valve) in September 2019 to 556 (40.4 per 1000 patients coded as having a mechanical valve) in May 2021. In the six months following the guidance, we observed a mean number of 557 (95% C.I = 538, 576), compared to a mean of 507 (95% C.I = 489, 525) in the six months prior. Charts of the monthly rate are available at [OpenSAFELY Reports](https://reports.opensafely.org/reports/doac-prescribing-in-people-with-a-mechanical-heart-valve-replacement/).

## Discussion

We identified 768 patients with codes suggesting the presence of mechanical heart valves that are currently prescribed DOACs, indicative of potentially inappropriate prescribing. We observed a progressive increase of potentially inappropriate prescribing between September 2019 and May 2021, with a 9.9% increase six months after the guidance was issued compared with six months before.

### Findings in context

In the UK there are approximately 10,000 valve replacement operations every year and in aortic valve replacement surgeries, a mechanical valve is used in approximately one in six^6,7^. We identified 15,457 people with a code for ever receiving a mechanical heart valve suggesting that not all mechanical valve replacements are explicitly coded in GP records. A US study showed an overall 1% rate of postoperative DOAC prescribing following surgical aortic or mitral valve replacement with mechanical valves between 2014 and 2017, in 18,000 people with comparable sex and age distributions to our study^8^. This is lower than the 6.8% rate observed in this study. We have previously described differences in the diagnosis of conditions and prescribing of certain medicines between GP clinical systems^9–12^. The reasons for the difference between EHR systems in this study is unclear but could be explained by true differences between EHR systems, the geographic variation in deployment of the systems^13^ and differences in the roll-out of the switching campaign at the height of a global health pandemic.

### Strengths and Weakness

The key strength of this study is the scale and completeness of the underlying raw EHR coded data, covering 97% of the English population. A limitation is in the comprehensive identification of people with a mechanical valve, as coded records may not explicitly state whether a valve is mechanical (e.g. *50733009 Replacement of mitral valve with prosthesis*) which may have led to under-ascertainment on the scale of the issue. We may also have counted a small number of people who were prescribed a DOAC before receiving a mechanical valve replacement, if it was fitted between March 2021 and May 2021. Finally, some people may be legitimately prescribed a DOAC whilst on a mechanical valve in rare clinical circumstances, with a justification recorded in the clinical record in free text. This data is not available but is likely to only account for a small number of all such patients.

### Policy Implications and future research

Anticoagulants are considered high risk in terms of patient safety^14,15^ and many national organisations have taken action to highlight specific clinical scenarios in breach of guidance through national safety alerts, cascaded via letters and e-mail^16,17^. For more targeted communication, using the analytical code in this report TPP and EMIS have directly alerted clinicians, through their EHR software, to draw attention to patients prescribed a DOAC who may have a mechanical heart valve. In addition, following an earlier version of this analysis NHS England issued a National Patient Safety Alert highlighting that clinicians should review affected patients. We will assess the impact of these two interventions in six months^18^.

This study demonstrates that a service with access to raw electronic health records data in near real time, such as OpenSAFELY, can be used to rapidly evaluate the impact of clinical guidance, and support rapid dissemination of audit and feedback via EHR software providers by sharing detailed code to identify specific classes of patient. We have made OpenSAFELY available to national NHS organisations for real-time monitoring and feedback, to measure and rapidly mitigate the direct and indirect health impacts of COVID-19 on the NHS

Further areas of investigation include: evaluating the impact of the rapid alert issued by EMIS and TPP, evaluating outcomes for those with a mechanical valve who may have inappropriately received a DOAC, and ascertaining the number of people for whom there is ambiguity on the type of valve they have received.

## Data Availability

All data were linked, stored, and analysed securely within the OpenSAFELY platform. Detailed pseudonymised patient data are potentially re-identifiable and therefore not shared. We rapidly delivered the OpenSAFELY data analysis platform without previous funding to deliver timely analyses of urgent research questions in the context of the global COVID-19 health emergency: now that the platform is established, we have established a process for external users to request access to the platform https://www.opensafely.org/onboarding-new-users/.

## Acknowledgements

We are very grateful for all the support received from the EMIS and TPP Technical Operations team throughout this work, and for generous assistance from the information governance and database teams at NHS England / NHSX, the national patient safety team at NHS England and NHS Improvement.

## Information governance and ethical approval

NHS England is the data controller; *EMIS and TPP are the data processors*; and the key researchers on OpenSAFELY are acting on behalf of NHS England. This implementation of OpenSAFELY is hosted within the *EMIS and TPP environments which are* accredited to the ISO 27001 information security standard and *are* NHS IG Toolkit compliant;^19,20^ patient data has been pseudonymised for analysis and linkage using industry standard cryptographic hashing techniques; all pseudonymised datasets transmitted for linkage onto OpenSAFELY are encrypted; access to the platform is via a virtual private network (VPN) connection, restricted to a small group of researchers; the researchers hold contracts with NHS England and only access the platform to initiate database queries and statistical models; all database activity is logged; only aggregate statistical outputs leave the platform environment following best practice for anonymisation of results such as statistical disclosure control for low cell counts.^21^ The OpenSAFELY research platform adheres to the obligations of the UK General Data Protection Regulation (GDPR) and the Data Protection Act 2018. In March 2020, the Secretary of State for Health and Social Care used powers under the UK Health Service (Control of Patient Information) Regulations 2002 (COPI) to require organisations to process confidential patient information for the purposes of protecting public health, providing healthcare services to the public and monitoring and managing the COVID-19 outbreak and incidents of exposure; this sets aside the requirement for patient consent.^22^ Taken together, these provide the legal bases to link patient datasets on the OpenSAFELY platform. GP practices, from which the primary care data are obtained, are required to share relevant health information to support the public health response to the pandemic, and have been informed of the OpenSAFELY analytics platform.

This study was approved by the Health Research Authority (REC reference 20/LO/0651) and by the LSHTM Ethics Board (reference 21863).

### Guarantor

BG/LS are guarantors of the OpenSAFELY project.

